# Non-Invasive Magnetic resonance imaging Biomarkers to evaLuatE histology-proven kidney fibrosis in Chronic Kidney Disease (NIMBLE-CKD): an observational cohort study protocol

**DOI:** 10.1101/2025.06.26.25330326

**Authors:** Dana Kim, Rebecca Kozor, Jason Chen, Muh Geot Wong, Martin Ugander

## Abstract

**Introduction:** Chronic kidney disease (CKD) affects 1 in 10 people worldwide and can progress towards kidney failure, which is best predicted by the severity of kidney fibrosis. Currently, kidney fibrosis can only be detected by invasive kidney biopsy which carries procedural risks with limitations on repeat testing. Magnetic resonance imaging (MRI) techniques have emerged as potential surrogate markers for kidney fibrosis, though data remains limited. To date, no studies have examined post-gadolinium contrast T1-mapping in kidney fibrosis despite its proven utility in assessing myocardial fibrosis. This study aims to develop a multiparametric MRI biomarker including post-contrast imaging to quantify kidney fibrosis in individuals with CKD.

**Methods and analysis:** In this observational cohort study, a control group of 20 healthy adult volunteers will establish healthy kidney MRI parameters. Two adult non-dialysis CKD cohorts (each n=24) who have undergone kidney biopsy within the last month will derive and validate the MRI models, respectively. Tubulointerstitial fibrosis on kidney biopsy will be assessed by Masson trichome staining and quantified based on the percentage of cortex affected by blinded pathologists. All participants will undergo a single multiparametric kidney MRI including kidney volumetry, T1- (pre- and post-low-dose contrast), T2- and T2*-mapping, diffusion weighted imaging and phase-contrast MRI of renal artery flow. The primary outcome will be the association between a composite multiparametric MRI marker and tubulointerstitial fibrosis with a minimum variance of 50%. The association between the multiparametric MRI marker and individual MRI variables, and tubulointerstitial fibrosis, estimated glomerular filtration rate and albuminuria will also be studied.

**Ethics and dissemination:** Ethics approval has been obtained by the Northern Sydney Local Health District Human Research Ethics Committee (2022/ETH00972). Results will be disseminated in relevant peer-reviewed journals and presented at academic conferences.

**Registration details:** This study is registered with the Australian New Zealand Clinical Trials Registry (ACTRN12622000855729p).

**STRENGTHS AND LIMITATIONS OF THIS STUDY:**

1. Kidney magnetic resonance imaging (MRI) is a growing area of interest due to its potential to provide a comprehensive assessment of kidney microstructure, haemodynamic properties, and function.
2. This study combines emerging and novel MRI techniques to develop a non-invasive biomarker for kidney fibrosis which can currently only be quantified histologically by kidney biopsy.
3. The multiparametric MRI models utilised in this study facilitates the examination of multiple MRI techniques including volumetry, T1-,T2-, and T2*-mapping, diffusion weighted imaging, and phase-contrast MRI in a single examination.
4. To our knowledge, this is the first study to examine post-contrast T1-mapping in kidney MRI in the detection of kidney fibrosis.
5. Limitations of this study include its limited sample size and lack of prospective follow-up, thus restricting its ability to report on the prognostic value of the proposed MRI biomarker.

## BACKGROUND

Chronic kidney disease (CKD) has a global prevalence of 13%, which is predicted to rise further^1^. It is an independent risk factor for morbidity and cardiovascular disease, and is a leading cause of non-communicable death worldwide^2^. As such, CKD has recently been recognised as a priority within the Word Health Organisation noncommunicable diseases agenda^3^. Kidney fibrosis is the strongest predictor and final hallmark of CKD regardless of underlying aetiology^4^. Early detection and targeted therapy is essential for preventing kidney failure yet routine clinical markers such as estimated glomerular filtration rate (eGFR) and proteinuria correlate poorly with kidney fibrosis, particularly in early disease^5^. Currently, the quantification of kidney fibrosis relies on invasive kidney biopsy, which is often performed at advanced stages when irreversible damage has already occurred. Biopsy carries risks with a high incidence of post-biopsy perinephric haematoma and pain^6^, making repeated procedures unsafe and preventing longitudinal monitoring of fibrosis. Further limitations of kidney biopsy include technical challenges based on patient anatomy and sampling error. Thus, there is a critical need for an alternative non-invasive biomarker to allow for the timely detection and monitoring of kidney fibrosis and identification of individuals at risk of kidney disease progression.

To this end, imaging modalities have emerged as promising tools in the assessment of CKD. In particular, there have been recent advancements in magnetic resonance imaging (MRI) techniques to assess renal function, microstructure, blood flow, perfusion and oxygenation^7-10^, which offer potential for earlier detection of CKD even before clinical manifestations appear. For example, total kidney volume measured from T1-or T2-weighted structural MR images is a validated biomarker in the assessment of kidney function, prognostication and treatment response in autosomal polycystic kidney disease, and has been shown to be associated with kidney function in the general population and other CKD aetiologies^7,11^. Techniques such as T1- and T2-mapping produce maps over the kidney to quantify tissue-specific T1- and T2-values respectively which reflect changes in the molecular environment^7^. T1-values are sensitive to water content, fibrosis and inflammation whereas T2-values are moreso affected by oedema and inflammation^6^, and there are a number of ongoing studies to assess their clinical applicability in CKD and kidney transplantation^12^. T2*-mapping utilises the paramagnetic properties of deoxygenated haemoglobin to indirectly assess tissue oxygenation^13^, and has been demonstrated to predict the risk of kidney function decline and kidney replacement therapy requirements in CKD^14^. Diffusion-weighted imaging (DWI) detects the displacement of water molecules in tissue, thus potentially aiding the assessment of fibrosis, perfusion, cellular infiltration and oedema in the kidney^15^. Phase-contrast (PC) MRI offers a non-invasive method to measure renal blood flow, enabling the assessment of renal haemodynamics in which changes occur at the earliest stages of kidney disease^16^.

A number of these MRI techniques have also been shown to correlate with histologically proven kidney fibrosis, with the most robust data being available for T1-mapping^17-19^, blood oxygen level dependent (BOLD) imaging derived from T2*-mapping,^20,21^ and DWI^17-20,22,23^. However, these findings have been inconsistent with limited sensitivity and specificity and are yet to be validated for clinical application, highlighting the need for further studies and development^9,24^. Multiparametric renal MRI protocols utilise multiple MRI techniques from a single examination to provide a comprehensive assessment of renal microstructure and function^25^. They are proposed to enhance the detection of kidney pathology and have been recommended to be applied in future studies rather than standalone techniques^7^. Indeed, multiparametric MRI models have shown to provide greater accuracy in predicting kidney fibrosis compared to single MRI measures^18^.

Importantly, studies of multiparametric renal MRI have avoided the use of gadolinium-based contrast agents (GBCA) to date due to concerns of developing nephrogenic systemic fibrosis (NSF) in CKD. However, a large meta-analysis has demonstrated that newer class II GBCA are safe even in advanced kidney disease^26^, and recent radiology guidelines no longer necessitate informed consent before administrating class II GBCA regardless of kidney function^27^. The use of post-contrast MRI has been of particular value in detecting myocardial fibrosis^28^ and is now routinely used in clinical practice. Specifically, areas of myocardial fibrosis are characterised by shorter T1 relaxation times following contrast administration^29^, and greater increases in 1/T1 (R1) from pre-to post-contrast, quantified as ΔR1, which is proportional to contrast agent concentration^30^. Additionally, post-contrast T1-mapping has been shown to correlate with fibrosis on myocardial biopsy^31^, making this an area with underexplored potential in renal pathology.

Here we outline an observational study protocol designed to explore several MRI techniques including novel methods utilising GBCA in multiparametric kidney MRI. The Non-Invasive Magnetic resonance imaging Biomarkers to evaLuatE histology proven kidney fibrosis in Chronic Kidney Disease (NIMBLE-CKD) study aims to develop a non-invasive imaging biomarker for kidney fibrosis in individuals with CKD.

## METHODS AND ANALYSIS

### Study design

The NIMBLE-CKD study is an observational cohort study that aims to develop a novel non-invasive MRI biomarker to predict the percentage of kidney fibrosis quantified by histology in individuals with CKD. A control group will consist of healthy volunteers to establish normal kidney MRI parameters. This cohort will not undergo kidney biopsy due to unacceptable risk in a healthy population. An MRI model will be derived from a cohort of individuals with CKD who have undergone kidney biopsy, which will validated against a second CKD participant cohort. The study cohorts, timeline and procedures are outlined in Figure 1.

**Figure 1.**
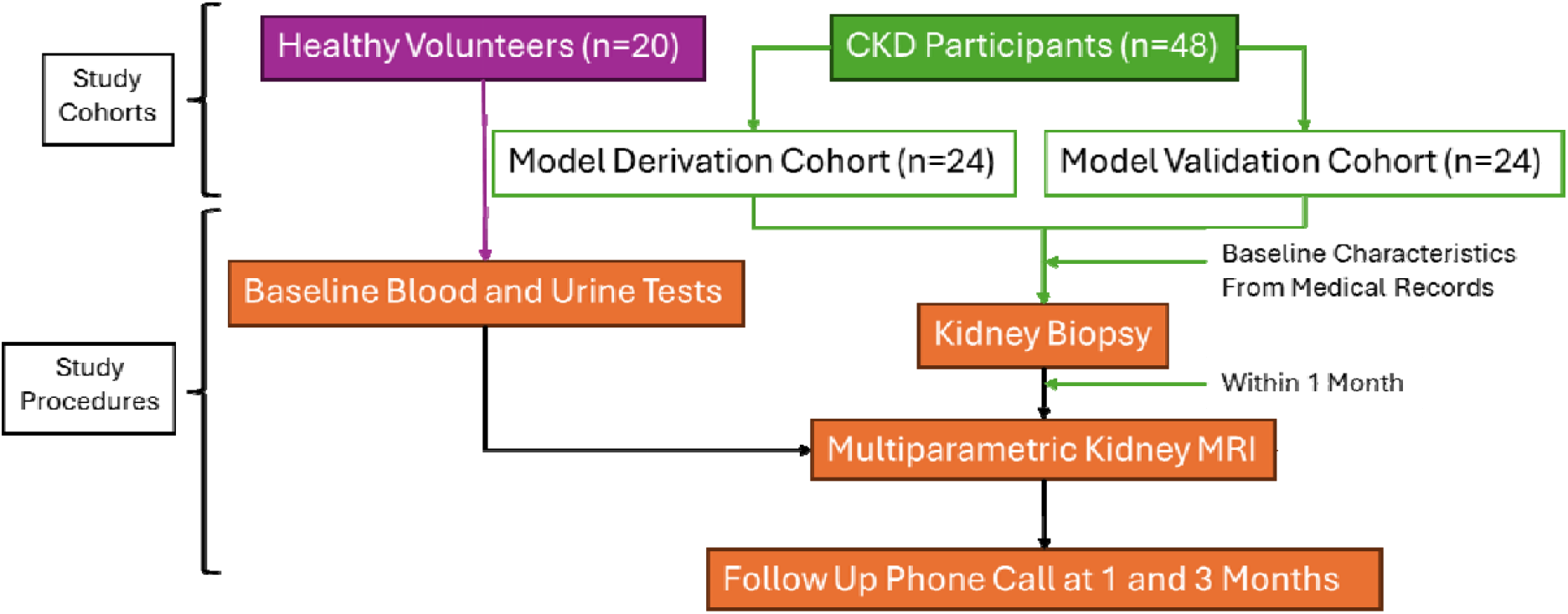
NIMBLE-CKD study cohorts and timeline. **Abbreviations:** CKD, chronic kidney disease; MRI, magnetic resonance imaging

### Study population

Healthy volunteers will be identified and recruited locally. Potential CKD participants planned for kidney biopsy due to clinical indications will be identified by their treating nephrologists. The inclusion criteria for healthy volunteers include age ≥18 years, no chronic medical conditions, estimated glomerular filtration rate (eGFR) >90mL/min/1.73m^2^ and absence of haematuria or proteinuria on urinalysis. The inclusion criteria for CKD participants include age ≥18 years, eGFR >15ml/min/1.73m^2^ and undergone kidney biopsy within one month. Exclusion criteria for all participants include a contraindication to MRI (e.g. MRI incompatible implantable devices, claustrophobia), contraindication to GBCA, and pregnant or breastfeeding females. Additional exclusion criteria for CKD participants include dialysis-dependent CKD and rapidly progressive glomerulonephritis on kidney biopsy.

### Data collection

#### Baseline characteristics

Baseline characteristics of all participants including age, height, weight, serum creatinine and eGFR based on the CKD-EPI formula^32^ will be collected and recorded. Additional clinical characteristics for CKD participants will be obtained from medical records including blood pressure, haemoglobin, urinary albumin excretion (spot albumin to creatinine ratio), presence of haematuria, medical comorbidities, and concomitant medications including diuretics, renin-angiotensin system inhibitors and sodium-glucose cotransporter-2 inhibitors.

#### Kidney biopsy and pathology analysis

Percutaneous kidney biopsy will be conducted using ultrasound or computed tomography-guidance based on the treating physician’s discretion. Histopathological samples of eligible participants will be reviewed prior to proceeding with MRI to ensure the exclusion criteria is not met.

All histopathological samples will be visually assessed by two independent renal pathologists, who will be blinded to the MRI results. Tubulointerstitial fibrosis (TIF) will be assessed by Masson trichome staining and quantified based on the percentage of cortex affected. The degree of tubulointerstitial fibrosis will be scored as a continuous variable from 0-100% and categorised based on the standard clinical scoring system (TIF 0 (<10%), TIF 1 (10-25%), TIF 2 (26-50%) or TIF 3 (>50%)). Other features of chronic kidney damage including glomerulosclerosis, tubular atrophy and arteriosclerosis will similarly be graded^33^. The presence of primary renal (glomerular, tubular, interstitial, vascular) pathology and the formal histopathological diagnosis will also be recorded.

#### Magnetic resonance imaging acquisition

All MRI will be acquired using a 3.0 Tesla MRI scanner with an 18-channel body array coil and an integrated 32-channel spine coil as the receiver (Siemens Healthcare, Erlangen, Germany). Multiparametric renal MRI (Figure 2) will be conducted over 45-minutes. Participants will be fasted for 4 hours and positioned supine with arms placed alongside the body.

**Figure 2.**
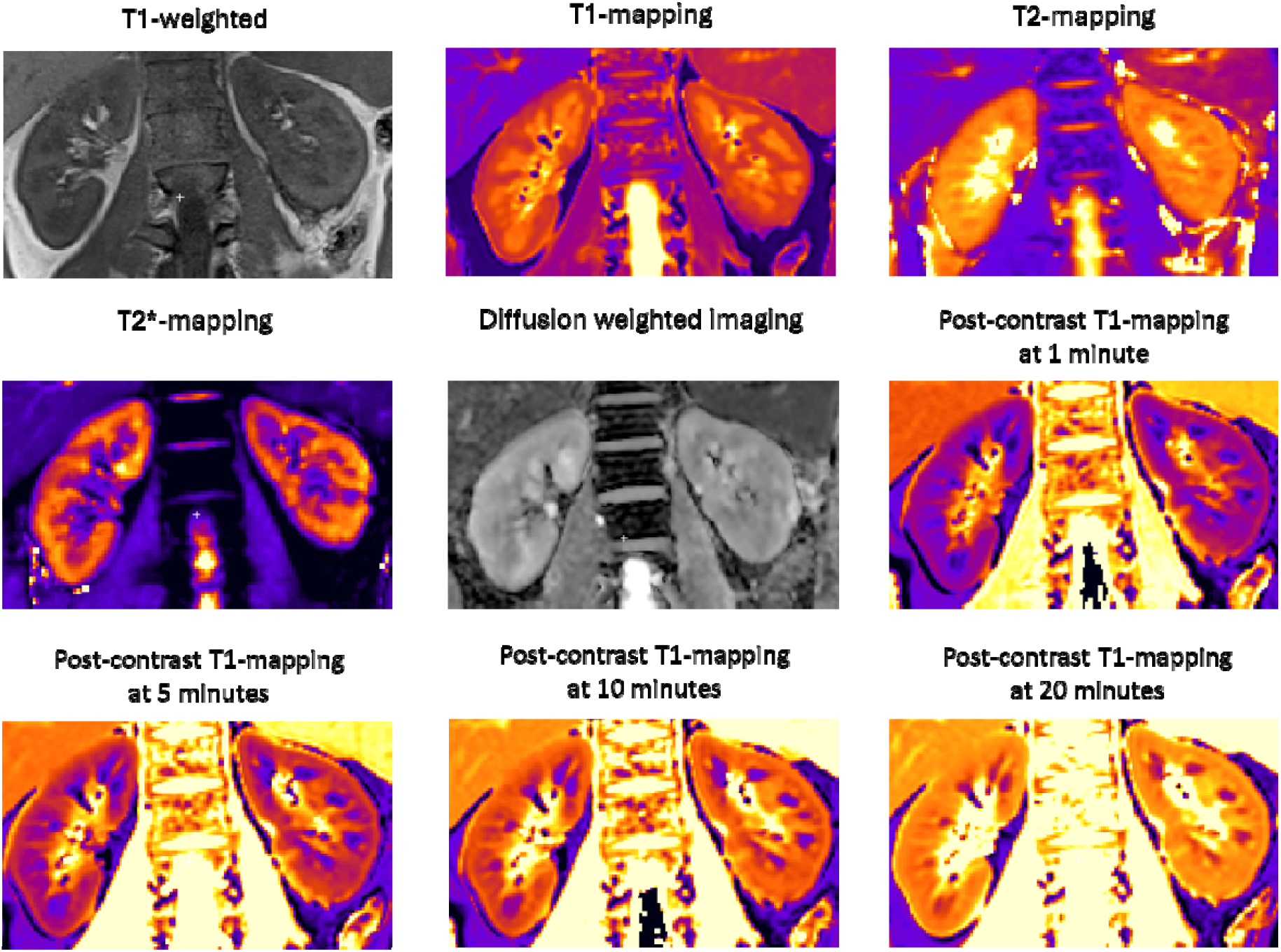
Representative multiparametric MR images acquired for the NIMBLE-CKD study. Example of different MRI techniques in the NIMBLE-CKD Study acquired in a chronic kidney disease participant.

Localisers in the axial, coronal and sagittal planes will be obtained for renal imaging. Coronal T1-weighted volumetric interpolated breath-hold examination (VIBE) sequences using a Dixon technique will be acquired to measure kidney volume. The acquisition parameters will include: slice thickness = 1.5mm, repetition time (TR) = 4.21 ms, echo times (TE) = 1.34 ms (out-of-phase) and 2.57 ms (in-phase), flip angle = 9°, field of view (FOV) = 260 × 320 mm, in-plane resolution = 1.25 × 1.25 mm^2^. Coronal T1-mapping will be performed using a modified Look-Locker inversion recovery (MOLLI) sequence with a 5(3)3 acquisition scheme during breath-hold. The parameters will include slice thickness = 8mm, TR = 401.68 ms, TE = 1.04 ms, flip angle = 35°, FOV = 218 × 256 mm, in-plane resolution = 1.48 × 1.48 mm^2^. Coronal T2-maps will be acquired using a T2-prepared steady-state free precession (SSFP) with the following parameters: slice thickness = 8mm, TR = 317.95 ms, TE = 1.25 ms (T2 Prep Duration 0, 35, 55 ms), flip angle = 12°, FOV = 154 × 192 mm, in-plane resolution = 1.98 × 1.98 mm^2^. Coronal T2*-maps will be acquired using a multi-echo gradient echo (GRE) sequence which will include the parameters: slice thickness = 8mm, TR = 54.95 ms, TE = 1.22, 2.36, 3.19, 4.30. 5.13 and 6.24 ms, flip angle = 20°, FOV = 156 × 192 mm, in-plane resolution = 1.98 × 1.98 mm^2^. Coronal diffusion-weighted imaging (DWI) will be performed using single-shot Echo Planar Imaging (ss-EPI) with Spectral Attenuated Inversion Recovery (SPAIR) fat suppression during free breathing with b-values of 50 and 800 s/mm^2^ and the following parameters: slice thickness = 8mm, TR = 3500 ms, TE = 68 ms, flip angle = 90°, FOV = 216 × 268 mm, in-plane resolution = 1.42 × 1.42 mm^2^. Phase contrast (PC) imaging will be performed in the through-plane perpendicular to the renal artery at the level of the hilum using a 2D gradient echo sequence with PC-velocity encoding (VENC) set at 100 cm/s. The imaging parameters will include: slice thickness = 6 mm, TR = 38.40 ms, TE = 2.65 ms, flip angle = 20°, FOV = 192 × 265 mm, in-plane resolution = 1.41 × 1.41 mm^2^.

Gadoteric acid, a class II macrocyclic agent, will be administered via a peripheral intravenous cannula at a dose of 0.05mmol/kg. No unconfounded cases of NSF have been reported with gadoteric acid^34^. Post-contrast T1-maps will be obtained at 1-, 5-, 10-, 15- and 20-minutes following contrast infusion utilising the T1-mapping parameters outlined above. All imaging acquisition parameters are detailed in Table 1.

**Table 1.** MRI techniques and acquisition parameters.

| Imaging Acquisition Parameter | MRI Technique |  |  |  |  |  |  |
| --- | --- | --- | --- | --- | --- | --- | --- |
|  | T1-Weighted | T1-Map | T2-Map | T2*-Map | DWI | Phase Contrast | Post-Contrast T1-Map |
| Sequence | VIBE | MOLLI 5(3)3 | T2-Prepared SSFP | Multiecho GRE | ss-EPI | PC-VENC GRE | MOLLI 5(3)3 |
| Orientation | Axial and coronal | Coronal | Coronal | Coronal | Coronal | Through-plane perpendicular to the renal artery | Coronal |
| Respiratory Control | Breath-hold | Breath-hold | Breath-hold | Breath-hold | Free-breathing | Breath-hold | Breath-hold |
| Slice Thickness (mm) | 1.5 | 8 | 8 | 8 | 8 | 6 | 8 |
| Slices (number) | 120 | 3 | 3 | 3 | 10 | 1 | 3 |
| TR (ms) | 4.21 | 401.68 | 317.95 | 54.95 | 3500 | 38.40 | 481.68 |
| TE (ms) | 1.34 (out of phase)<br>2.57 (in-phase) | 1.04 | 1.25<br>T2 Prep Duration<br>0, 35, 55 | 1.22, 2.36,<br>3.19, 4.30,<br>5.13, 6.24 | 68 | 2.65 | 1.04 |
| Flip Angle (°) | 9 | 35 | 12 | 20 | 90 | 20 | 35 |
| FOV (mm) | 260 x 320 | 218 x 256 | 154 x 192 | 156 x 192 | 216 x 268 | 192 x 265 | 218 x 256 |
| In-plane Resolution (mm) | 1.25 x 1.25 | 1.48 x 1.48 | 1.98 x 1.98 | 1.98 x 1.98 | 1.42 x 1.42 | 1.41 x 1.41 | 1.48 x 1.48 |

**Abbreviations:** DWI, diffusion weighted imaging; TR, repetition time; TE, echo time; FOV, field of view; VIBE, volumetric interpolated breath-hold examination; MOLLI, modified Look-Locker inversion recovery; SSFP, Steady-State Free Precession; GRE, gradient echo; ss-EPI, single-shot Echo Planar Imaging; PC-VENC, phase-contrast velocity encoded segmented

#### Magnetic resonance imaging analysis

MR images will be analysed using the image analysis software Segment (Medviso, Lund, Sweden) by two independent investigators blinded to the participant’s baseline laboratory and biopsy results. All analyses will be repeated for the left and right kidneys for each participant. Total kidney, cortical and medullary volume will be measured by manually placing regions of interest (ROIs) on each slice of the coronal T1-weighted VIBE in-phase images. Motion corrected images will be used to measure T1, T2, T2* and post-contrast T1 values. Apparent diffusion coefficient (ADC) maps will be generated from DWI. A single ROI will be manually drawn on the T1, T2, T2*, post-contrast T1- and ADC maps in the kidney cortex and medulla, avoiding areas of heterogeneity, vascular structures, cysts, haemorrhage, and artifacts. Cortical and medullary values will be reported, alongside the cortical-medullary difference and cortical-medullary ratio. Cortical and medullary contrast distribution will be quantified from post-contrast T1-mapping images over time. Renal artery blood flow analysis will be performed using phase-contrast scans with static tissue background correction. A manual ROI will be placed on the renal artery across all phases of the cardiac cycle. Renal perfusion will be calculated by dividing the mean renal artery blood flow by total kidney volume.

### Outcomes

The primary outcome will be the association between a multiparametric MRI marker and tubulointerstitial fibrosis quantified on kidney biopsy with a minimum variance of 50% (R^2^=0.5). The model for the primary outcome will include the following MRI variables: total kidney volume, cortical T1 value, cortical T2 value, cortical T2* value, cortical ADC value, renal artery blood flow, renal perfusion, and cortical contrast distribution measures.

The secondary outcomes will include the association between 1) individual MRI variables and tubulointerstitial fibrosis, 2) combinations of MRI variables and tubulointerstitial fibrosis, based on significant correlations identified in 1), 3) the multiparametric MRI marker for the primary outcome and eGFR and 4) the multiparametric MRI marker for the primary outcome and albuminuria. Exploratory outcomes will include the association between 1) the multiparametric MRI marker for the primary outcome and tubulointerstitial fibrosis as a categorical variable, and 2) individual MRI variables and tubulointerstitial fibrosis as a categorical variable.

### Sample size calculation

There will be 20 participants in the healthy volunteer cohort. To detect a minimum of 50% variance explained by eight MRI variables, at a 0.05 level of significance and minimum power of 0.80, a sample of 24 subjects is required. Therefore, there will be 24 participants in each of the CKD participant derivation and validation cohorts.

### Data analysis plan

All statistical analyses will be performed using R Statistical Software (v4.1.3; R Core Team 2022). Continuous variables will be expressed as mean and standard deviation and compared using t-tests for normally distributed data. Skewed data will be expressed as median and interquartile range and compared using Mann-Whitney U tests. Data normality will be assessed visually using histograms. Categorical data will be compared using chi-squared tests. The healthy control group will determine normal MRI reference ranges for baseline comparison. All variables (MRI, tubulointerstitial fibrosis, eGFR and albuminuria) will be measured and reported as continuous variables for the primary and secondary outcomes. Tubulointerstitial fibrosis will be reported as a 4-level categorical variable for the exploratory outcomes. Uni- and multivariable linear regression models will be fitted to examine the association between the multiparametric MRI model and individual MRI markers with tubulointerstitial fibrosis, eGFR and albuminuria. T-tests and analysis of variance (ANOVA) will be used to test the significance of the models. The models will be tested and developed in the CKD participant derivation cohort. A second CKD participant validation cohort will be used to evaluate the accuracy and sensitivity of all significant models. All statistical tests will be two-sided and performed at a 0.05 level of significance.

### Safety monitoring

All serious adverse events that occur during the study will be documented by the Investigator and reported to the relevant governing bodies. Additional reportable GBCA-associated adverse events include serious allergic-like reactions requiring treatment or hospitalisation, and nephrogenic systemic fibrosis within 3-months of contrast administration^35^. Complications related to kidney biopsy will not be documented for the purpose of this study as this procedure is within the scope of standard clinical practice and will be managed accordingly by the participant’s treating nephrologist.

### Patient and public involvement

The George Institute for Global Health Consumer Kidney Panel was involved in the review of the study methodology, procedures, and patient information consent form.

### Study status

Participant recruitment commenced in July 2024 and is ongoing.

### Ethics and dissemination

This study has been approved by the Northern Sydney Local Health District Human Research Ethics Committee (2022/ETH00972), with governance approval from Royal North Shore Hospital (2022/STE01742) and New South Wales Health Pathology (2023/STE03679) and is registered with the Australian New Zealand Clinical Trials Registry (ACTRN12622000855729p). Results will be disseminated in relevant peer-reviewed journals and presented at academic conferences. All manuscripts will be reviewed and approved by all investigators prior to publication. Authorship will be based on contribution to the manuscript.

## DISCUSSION

Early detection of CKD and identification of those at risk of kidney failure is important to guide prognostication and treatment decisions. Current clinical markers for estimating kidney function such as eGFR are notoriously fraught with limitations^36^ and the assessment of kidney fibrosis by kidney biopsy comes with risk. Here we propose to develop a non-invasive imaging biomarker for histology quantified kidney fibrosis and explore its relationship with eGFR and albuminuria. By utilising multiparametric models, we aim to build upon existing literature for MRI techniques including T1-mapping, T2*-mapping and DWI and examine emerging techniques such as PC-MRI. We will also provide novel data on the application of post-contrast T1-mapping in kidney MRI and kidney fibrosis which, to the best of our knowledge, has not been previously investigated. By leveraging established techniques in cardiac MRI, the addition of post-contrast imaging alongside previously studied kidney MRI techniques has the potential to profoundly enrich the assessment of kidney fibrosis in CKD.

Limitations including cost, accessibility and inconsistent or limited data have hindered the clinical application of kidney MRI biomarkers to date^37^. Global collaborative efforts are underway to standardise pre-clinical kidney MRI protocols^8,38^ yet knowledge gaps persist. For example, experts have failed to reach consensus on factors such as the impact of medication and hydration or fasting status on MR images, and image analysis techniques^39,40^. Limitations specific to our study include the limited sample size and the use of a single MRI scanner which may preclude direct application of our MRI protocols to other scanners. Furthermore, as we will be acquiring a single MRI examination per participant, we will be unable to assess disease progression or report on the prognostic value of these MRI models at this stage.

Despite these limitations, kidney MRI remains an evolving field of great interest due to its ability to provide a wide breadth of information regarding kidney microstructure, haemodynamic properties, and function. Whilst multiple imaging techniques have been studied in CKD, MRI has the most data in detecting kidney fibrosis such that it is the only modality to be utilised as an imaging endpoint in clinical trials^41^. Certainly, the non-invasive nature of MRI and ability to be safely repeated over time makes it attractive for longitudinal monitoring of disease. In addition, with the focus on anti-fibrotic therapies in the prevention of CKD progression^42,43^, the conceivable use of MRI to assess efficacy in reducing kidney fibrosis to further drug-development is highly anticipated. The NIMBLE-CKD study aims to advance our ability to assess kidney fibrosis and serve as a platform for larger prospective studies to ultimately demonstrate the translational potential of multiparametric kidney MRI biomarkers for kidney failure risk stratification in clinical practice and surrogate kidney outcomes in clinical trials.

## Data Availability

All data produced in the present study are available upon reasonable request to the authors.

## AUTHORS’ CONTRIBUTIONS

MGW and MU conceived the study. All authors contributed towards the design of the study. DK drafted the manuscript and RK, JC, MGW and MU revised it critically for important intellectual content. All authors approved of the final version to be published.

## FUNDING STATEMENT

This study received start-up academic funding from the University of Sydney.

## ACKNOWLEDGEMENTS

DK was supported by a Jacquot Research Entry Scholarship from the Royal Australian College of Physicians Foundation for this study. This research was supported by New South Wales Health Pathology through reviews of kidney biopsy samples.

## COMPETING INTERESTS STATEMENT

DK reports no conflicts of interest. MGW has received fees for advisory boards, steering committee roles, or scientific presentations from Travere, Baxter, Amgen, Abbvie, Chinook, Dimerix, Ostuka, GlaxoSmithKline and CSL-Behring. MU is affiliated with Karolinska Institute and the University of Sydney both of which have research and development agreements regarding cardiovascular MRI with Siemens.

## Notes

### Author Declarations

The Human Research Ethics Committee of the Northern Sydney Local Health District gave ethical approval for this work (2022/ETH00972).

### Summary of Updates

Minor revisions to include the recent inclusion of chronic kidney disease as a WHO priority and to clarify the methodology within the abstract.

